# Characterization of CXCL10 as a biomarker of respiratory tract infections detectable by open-source lateral flow immunoassay

**DOI:** 10.1101/2024.01.12.24301261

**Authors:** Dayna Mikkelsen, Jennifer A. Aguiar, Benjamin J-M Tremblay, Manjot S. Hunjan, Ulrich Eckhard, Jodi Gilchrist, David Bulir, Marek Smieja, Samira Mubareka, Catherine Lambert, Kha Tram, Andrew C. Doxey, Jeremy A. Hirota

## Abstract

Understanding core mechanisms common to respiratory tract viral pathogenesis and host-responses to infections may provide biomarkers for at-risk patient populations that guide interventions aimed at reducing morbidity, mortality, and economic costs. Secreted interferon stimulated gene protein products including CXCL10, CXCL11, and TNFSF10 could provide early biomarker signals that are prognostic for respiratory tract viral infections. In the present study, we had the overarching goal of defining the expression patterns of CXCL10, CXCL11, and TNFSF10 in clinical respiratory mucosal samples for multiple respiratory tract infections including respiratory syncytial virus, rhinovirus, influenza A and SARS-CoV-2 to inform the development of a host-biomarker point of care lateral flow immunoassay tool.

Gene expression levels from upper airway samples suggested that *CXCL10* and *CXCL11* elevations were consistent across multiple viruses, correlated with higher SARS-CoV-2 viral load, and had a lower variance over the course of COVID-19 infection compared to *TNFSF10*. Deep proteomic profiling using mass-spectrometry revealed CXCL10 protein was not detectable in oral samples from healthy individuals. CXCL10 levels were measured from the saliva of SARS-CoV-2 infected individuals and showed significant elevations in CXCL10 protein concentration. A prototype lateral flow immunoassay for detecting CXCL10 protein with a sensitivity of 2ng/mL in human saliva is presented.

Our work provides a foundation for further exploration of CXCL10 as a host biomarker relevant in respiratory tract viral infections. Leveraging lateral flow immunoassay technology for detection of biomarkers prognostic of respiratory tract infection may provide opportunities to intervene selectively and aggressively in those most at risk of poor outcomes.

## Introduction

Prior to COVID-19, lower respiratory infections ranked as the 4^th^ leading cause of death responsible for ∼ 2.6 million fatalities annually [1]. The majority of these deaths occurred in children under the age of five and those over the age of 70[1]. The COVID-19 pandemic exacerbated the global burden of disease attributable to respiratory infections with more than 6 million fatalities [2] where age functioned as a significant risk factor that has spared children from the greatest morbidity and mortality [3].

The morbidity and mortality associated with respiratory tract infections are dominant aspects that contribute to a global burden calculation, but these measures must also be considered in conjunction with the economic costs [4]. Economic costs of respiratory infections can be broken down into direct and indirect costs. Direct costs of respiratory infections are incurred through interactions with the healthcare system and may include physician visits and medication costs. Indirect costs may include absenteeism for patients and caregivers [5]. Total costs of pneumonia and acute lower respiratory infections exceed $25 billion annually in the European Union for a population of ∼450 million people[6]. Extrapolating these costs on a global population level estimates ∼$400 billion in annual costs across all jurisdictions. When including COVID-19, these expenditures are much higher. On a global scale, the economic burden of COVID-19 led to a -3.1 % decrease (equal to -$2.4 trillion USD) in the annual global GDP [7], with total estimates potentially as high as $16 trillion USD[8]. The COVID-19 pandemic demonstrated that in dire circumstances, respiratory infections can cause unprecedented global economic recession and massive disruptions to global supply chains[9]. While the economy has begun to recover from the COVID-19 pandemic, the risk of novel emerging infectious diseases and SARS-CoV-2 variants of concern remains high. The significant direct and indirect costs of respiratory infections warrant the development of a diverse set of approaches that can help mitigate economic burden.

Across common respiratory tract viral infections, the young, elderly, individuals with chronic respiratory diseases, and expecting mothers have elevated risks for morbidity and mortality [10]. Common viral respiratory tract pathogens include rhinoviruses (RV), respiratory syncytial virus (RSV), influenza A and B (FluA and FluB), and coronaviruses (CoVs) [3,11]. Depending on the pathogen, respiratory viral infections may vary in presentation from a mild illness restricted to upper airways that is self-resolving (RV) [12] to acute respiratory distress syndrome that requires ventilation and is associated with fatalities (SARS-CoV-2) [3]. Globally it has been estimated that 2% of all deaths related to respiratory dysfunction are associated with influenza causing approximately ∼400 000 deaths every year [13]. RV cause relatively mild illness in the majority of the population; although they contribute to a substantial number of medical visits and missed work days every year and an increased morbidity is observed in immunocompromised individuals [14]. RV infections contribute to a substantial number of acute asthma exacerbations in the community [15]. RSV is a common viral infection in children under 5 years old that is associated with hospitalizations and bronchiolitis [16,17]. In a longitudinal study of 92 children, all but one had an RSV infection before the age of 2, suggesting that a large proportion of children are susceptible to becoming infected with RSV [18]. In 2015, there were approximately 33 million RSV infections globally which resulted in three million hospitalizations and ∼ 60,000 deaths in children younger than 5 years old [19]. Both RV and RSV infections pose as risk factors for subsequent development of wheeze and childhood asthma [20,21]. Understanding core mechanisms common to respiratory tract viral pathogenesis and host-responses to infections may provide biomarkers for at-risk patient populations that guide interventions aimed at reducing morbidity, mortality, and economic costs.

To combat respiratory tract viral infections, the respiratory mucosa participates in a wide range of innate and adaptive immune responses to control and eliminate pathogens. Innate immune responses of the respiratory mucosa are initiated by germ-line encoded pattern recognition receptors (PRRs) that bind to molecular motifs that are associated with extrinsic pathogen associated molecular patterns (PAMPS) or intrinsic damage associated molecular patterns (DAMPS). PRR recognition of non-self PAMPS or self-DAMPS initiates intracellular signaling cascades that upregulate host-defence mechanisms defined by temporally specific gene expression patterns [22]. Although these processes are broadly applicable to all respiratory tract viral infections, diversity in host responses exist and are influenced by the infectious pathogen and its ability to subvert host immune responses in an attempt to replicate and spread [23,24]. Core host innate immune responses that are shared between respiratory tract viral infections may provide diagnostic and prognostic potential beyond simple pathogen detection alone [25,26]. Importantly, host innate immune responses are not merely beneficial and can contribute to pulmonary damage via uncontrolled inflammation and immunopathology [27].

The dominant innate immune response induced upon respiratory tract viral infection and downstream of PRR signaling is the induction of interferon stimulated genes (ISGs) through Type I and Type III interferons [24]. When a virus enters the respiratory system, viral PAMPs are recognized by the host’s PRRs, and this triggers the production of interferons (type I and III) and activates the interferon antiviral response. Type I IFNs act in a paracrine and endocrine fashion through the Type I IFN receptor (IFNAR) while Type III IFNs signal through IFNLR1 [24]. Downstream signaling from these receptors merges on the JAK family of kinases and STAT family of transcription factors, leading to transcriptional activation[28]. The interferon antiviral response induces a number of different ISGs that generate protein products that inhibit distinct parts of viral life cycles [24,28]. Aspects that the interferon antiviral response may coordinate include cellular defences to virus binding, internalization, replication, packaging, and release. Importantly, in conjunction with ISG products produced *within* the cell to combat viral life cycle, other ISG products are released *outside* the cell to signal to non-infected cells in the local and systemic environment. Secreted ISG protein products that are also chemokines may provide early biomarker signals that are prognostic for respiratory tract viral infections. Three of these chemokines include Tumour Necrosis Factor-related Apoptosis-Inducing Ligand (TRAIL) and the CXC Chemokines: CXCL10 and CXCL11 which are all induced by interferon [23]. Previous research has found these three proteins to be induced by several respiratory viral infections both *in-vitro* and *in-vivo* experiments [29–31]. Together, these three genes *CXCL10*, *CXCL11* and *TNFSF10* are cytokines that are also stimulated by the IFN response and may be useful biomarkers for respiratory tract viral infections.

CXCL10, also called interferon-γ inducible protein 10 kDa (IP-10), is a secreted ISG and plays a key role in fighting viral infections [32]. This chemokine has been shown to have altered mRNA expression levels in RV, RSV, and SARS-CoV-2 infections [33–35]. In addition, serum samples revealed high levels of CXCL10 protein in H5N1 influenza infection and SARS-CoV-2 [36,37]. Lorè *et al.* 2021 showed CXCL10 serum levels were a robust predictor of COVID-19 outcomes, with higher levels being related to increased mortality. CXCL11, also known as C-X-C motif chemokine ligand 11, is another chemokine which shares the same CXCR3 receptor as CXCL10 and is also secreted[38]. Both CXCL10 and CXCL11 play a role in recruitment of T cells and NK cells as part of the immune response [38]. CXCL11 has shown a similar trend of upregulation in SARS-CoV-2 infections *in-vitro* [39] and *in-vivo* [35,40]. TRAIL produced from the gene *TNFSF10*, is a secreted protein which can induce apoptosis and plays a role in the immune response to destroy virally infected cells [41]. *TNFSF10* has been found to be upregulated during viral infection [42].

Respiratory tract infections caused by viruses have sought a massive burden on health care system for years and the recent COVID-19 pandemic has exacerbated this exponentially. The COVID-19 pandemic has ushered in a new era of rapid diagnostics, including the use of lateral flow assays (LFAs) in settings other than hospitals, such as schools and workplaces. LFAs are useful for testing directly on-site rather than operating through a centralized testing laboratory. The advantages of LFA tests are that they are quick, inexpensive, and easy to use. Despite their lower sensitivity and specificity, they are typically preferred as a screening method and require a confirmation test such as PCR [43]. SARS-CoV-2 rapid tests such as the Abbott Panbio™ detect the SARS-CoV-2 pathogen but do not provide any insight into the individual’s host response and disease trajectory. However, orthogonal tests, such as Roche’s Elecsys® IL-6 test, which employs an electrochemiluminescence immunoassay (ECLIA) to measure IL-6 levels in blood, have demonstrated promising use for the early detection of severe disease progressions in COVID-19 patients [44]. Leveraging LFA technology for detection of biomarkers prognostic of respiratory tract infection may provide opportunities to intervene selectively and aggressively in those most at risk of poor outcomes.

ISGs play a major role in the antiviral defense and measuring extracellular protein products of ISGs could be potential biomarkers of infection. CXCL10, CXCL11 and TNFSF10 are involved in the antiviral immune response during a variety of respiratory infections. However, the feasibility and performance of these ISGs as useful and measurable clinical biomarkers from an easily accessible sample format remains unknown. In the present study, we had the overarching goal of defining the expression patterns of the secreted ISG products CXCL10, CXCL11, and TNFSF10 in respiratory mucosal samples for multiple respiratory tract infections including RSV, RV, influenza A and SARS-CoV-2 to inform the development of a host-biomarker point of care LFA tool. Gene expression levels from upper airway samples suggested that *CXCL10* and *CXCL11* elevations were consistent across multiple viruses, correlated with higher SARS-CoV-2 viral load, and had a lower variance over the course of COVID-19 infection compared to *TNFSF10*. Deep proteomic profiling using mass-spectrometry revealed CXCL10 protein was not detectable in oral samples from healthy individuals. CXCL10 levels were measured from the saliva of SARS-CoV-2 infected individuals and showed significant elevations in CXCL10 protein concentration. We proceeded to develop a prototype LFA for CXCL10 protein with a sensitivity of 2ng/mL in human saliva. Our work provides a foundation for further exploration of CXCL10 as a host biomarker relevant in respiratory tract viral infections with potential diagnostic and prognostic value.

## Materials and Methods

### Human ethics

Procurement of nasopharyngeal swab (NPS) and saliva samples from consented study subjects was approved by Hamilton integrated Research Ethics Board (HiREB 4914T, 5099T, and 10771).

### Data Resources

A combination of publicly available gene and protein datasets (**Table 1**) and in-house generated gene datasets from Hamilton Regional Laboratory Medicine Program and Sunnybrook Hospital were used for present study.

**Table 1:**
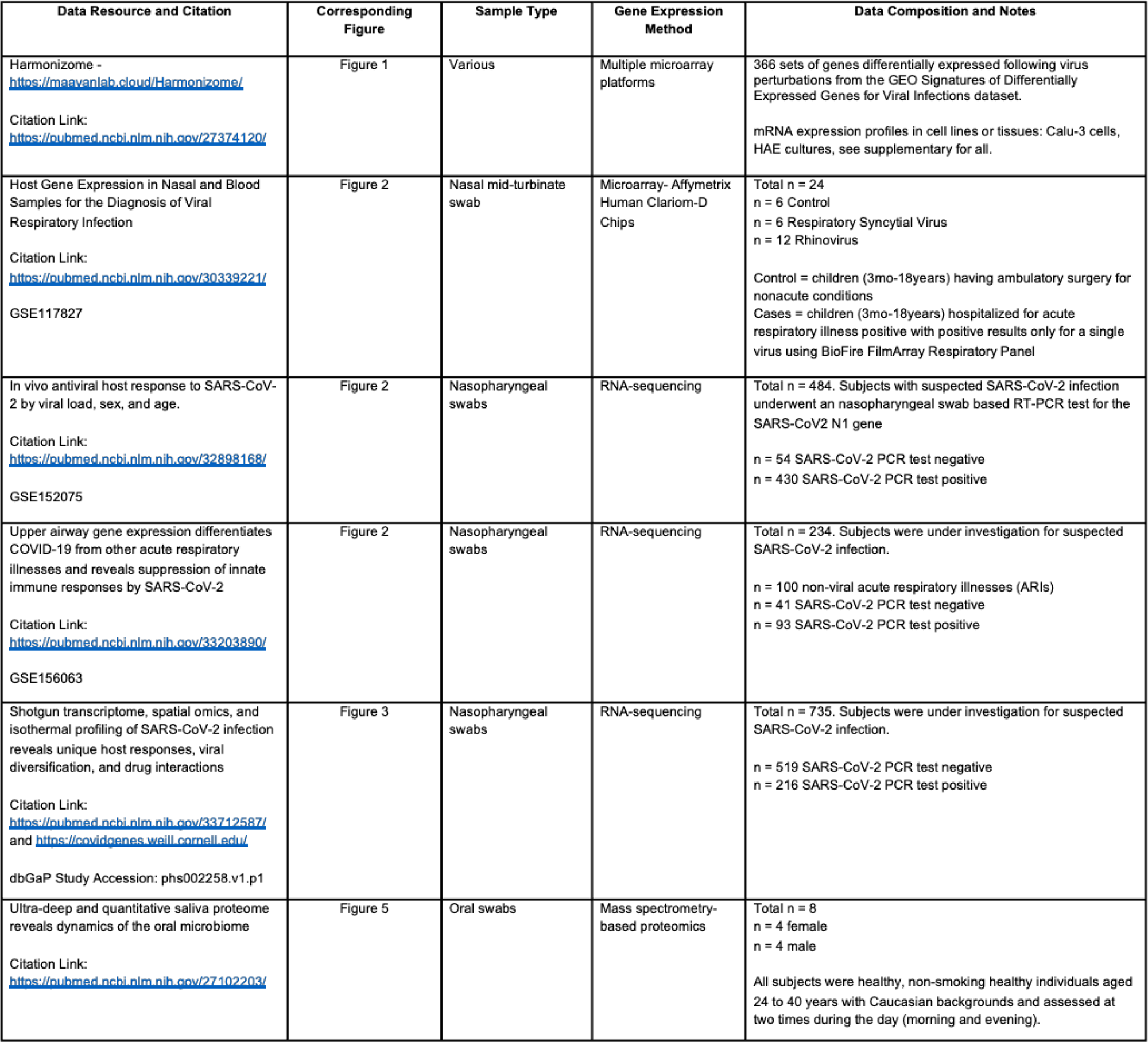
Publicly Available Data Resources Accessed for Analysis.

### Overview of Publicly Available Accessed Datasets

#### *In-vitro* cell culture experiments

*In-vitro*, pre-processed transcriptomic datasets were examined using the Harmonizome resource (https://maayanlab.cloud/Harmonizome/ [45]. From Harmonizome, the dataset labelled “GEO Signatures of Differentially Expressed Genes for Viral Infections” was accessed for hypothesis testing for differential expression of in *CXCL10*, *CXCL11*, and *TNFSF10* following viral exposure *in vitro* from a variety of experimental conditions. For each dataset, Harmonizome’s “Standard Value” reports the strength of association as a -log_10_(*p*-value) with the sign indicating up (+) or down (-) expression, respectively. The “GEO Signatures of Differentially Expressed Genes for Viral Infections” dataset contained 366 individual datasets of mRNA expression profiles using microarray technology for cell lines or tissues after viral infection. These were filtered down to 199 microarray datasets as all non-human experiments and non-respiratory viruses were excluded. The remaining 199 datasets came from 17 GSE studies including the following viruses: influenza A, human coronavirus, SARS-CoV like virus isolated from bats, a mouse-adapted SARS-coronavirus, SARS-CoV mutant strain that does not express the accessory protein open reading frame 6 (delta ORF6), human metapneumovirus, wild-type infectious clone-derived SARS-CoV, measles, RSV and RV, with time points ranging from 0-96 hours post inoculation. *In-vitro* cell systems included but were not limited to Calu-3, HAE, and A549 cell lines (Supplementary Table 1). From the 199 microarray datasets, the studies in which *CXCL10, CXCL11* and *TNFSF10* were in the top 300 or bottom 300 differentially expressed were counted. The Standard Value (strength of association) and number of datasets were plotted. This Harmonizome method of data retrieval is a conservative representation of the number of datasets where the candidate genes are differentially regulated as they must be in the top or bottom 300 differentially expressed genes. *ACTB*, *TUBB* and *GADPH* were analyzed as housekeeping genes in parallel to demonstrate the stochastic nature intrinsic to the approach used for our genes of interest.

#### Upper Airway Sampling

Four datasets of upper airway swabs (NPS or nasal mid-turbinate) samples were curated to determine *CXCL10*, *CXCL11* and *TNFSF10* gene expression under different respiratory viral infections.

Host transcriptomic data on RV and RSV infections were from pediatric populations between 3 months and 18 years admitted to hospital for acute respiratory illness and had confirmed PCR detection of pathogen [46]. All subjects had not received any immunosuppressants for 30 days or antibiotics for 7 days. Control subjects were of the same age but present in the hospital setting for ambulatory surgery for non-acute conditions. Microarray gene expression analysis was performed on the mid-turbinate nasal swab with data retrieved from GSE117827. A total of 6 RSV positive (2 males, 4 females), 12 RV positive (8 males and 4 females) and 6 negative control (6 males) samples were analyzed.

Host transcriptomic data on SARS-CoV-2 infections were from adult populations from three studies that each had distinct subject enrolment strategies and demographics available. In the first dataset, a total of 430 SARS-CoV-2 positive (176 males, 201 females, 53 unknown) and 54 negative control (30 males, 24 females) were analyzed from GSE152075 [35]. For the second dataset, a total of 93 SARS-CoV-2 positive (no other pathogenic respiratory virus) (50 females, 43 unknown), 41 other viral acute respiratory infection (19 females, 22 unknown), and 100 non-viral acute respiratory infection (55 females, 45 unknown) were analyzed from GSE156063 [47]. For the third dataset, SARS-CoV-2 status was determined by RT-PCR and other respiratory viruses were detected by metagenomic sequencing. The non-viral acute respiratory infection group included bacterial infections and other non-infectious respiratory diseases. A total of 216 SARS-CoV-2 and 519 negative samples were analyzed from Genotypes and Phenotypes dbGAP (accession #38851 and ID phs002258.v1.p1) [40].

To determine the association between positive SARS-CoV-2 status and *CXCL10, CXCL11* and *TNFSF10* expression, GSE152075 and GSE156063 were accessed. To determine the association between SARS-CoV-2 viral load and *CXCL10, CXCL11* and *TNFSF10* expression a web portal for an RNA-sequencing dataset was accessed (https://covidgenes.weill.cornell.edu/). We extracted the log_2_ fold change and q values for *CXCL10, CXCL11* and *TNFSF10* for positive vs negative comparison group and “Viral Level Continuous” comparison group using SVA correction and included all reads. “Viral Level Continuous” comparison group converted SARS-CoV-2 qRT-PCR cycle threshold (Ct) values into a continuous variable by converting Ct values where Ct of 15 is equal to 1.0 and a Ct over 40 is taken as 0.0 (see - https://covidgenes.weill.cornell.edu/). The goal of this was to determine if *CXCL10, CXC11* or *TNFSF10* gene expression correlated with SARS-CoV-2 viral load.

#### Oral Sampling

To determine the expression level of CXCL10, CXCL11, and TNFSF10 at the protein level in healthy individuals, an ultra-deep liquid chromatography-mass spectrometry-based analysis of the human saliva proteome was accessed that included 8 subjects (4 males and 4 females) (PXD 003028) [48]. All subjects were healthy and asymptomatic with no oral inflammation, pathologies, or prescribed medications. Each subject was sampled twice, once in the morning and once after breakfast.

### Overview of Analysis of In-House Datasets

#### Upper Airway Sampling

Three datasets of NPS upper airway samples were generated to determine *CXCL10*, *CXCL11* and *TNFSF10* gene expression under influenza A and SARS-CoV-2 infection. For influenza A infections, NPS samples from patients suspected of respiratory tract infection were collected in the first 6 months of 2020 through the Hamilton Regional Laboratory Medicine Program (HRLMP) during COVID-19 screening. NPS swabs (Copan, Italia) were collected and stored in universal transport media prior to multiplex PCR analysis for a panel of respiratory tract infections. A total of 8 influenza A positive (5 males, 3 females) and 14 negative control (7 males, 7 females) samples were analyzed. For comparison of fatal and non-fatal SARS-CoV-2 infections, NPS samples from positive for COVID-19 were collected in the first 6 months of 2020 through the Sunnybrook Hospital during regional COVID-19 testing programs. NPS swabs (Copan, Italia) were collected and stored in universal transport media prior to RNA isolation. A total of 6 fatal (5 males, 1 female) and 19 non-fatal (9 males, 10 females) samples were analyzed. For determining the variability of *CXCL10, CXCL11*, and *TNFSF10* over time following a positive SARS-CoV-2 test, PCR confirmed COVID-19 positive patients underwent repeated NPS sampling followed by storage in McMaster Molecular Medium and RNA isolation. A total of n = 6 subjects (5 females, 1 male) were analyzed over the period in which each subject was an inpatient (range from 12 to 26 days). All RNA isolation was performed using New England Biolabs Monarch Total RNA Miniprep Kit according to manufacturer directions. RNA concentration and integrity number was assessed by Agilent Bioanalzyer according to the manufacturer’s directions. Gene expression profiling was performed using the Human Clariom D microarray assay (Applied Biosystems).

#### Oral sampling

To determine the suitability for measuring CXCL10 protein in saliva as a surrogate for *CXCL10* gene in upper airway samples, we assessed CXCL10 protein by multiplex cytokine array (Eve Technologies, Calgary, Alberta) in COVID-19 inpatients processed by the Infectious Disease Research Group at Research St. Joseph’s - Hamilton. Neat saliva samples were provided from SARS-CoV-2 positive inpatients (n = 3) and SARS-CoV-2 negative healthy controls (n = 6). Samples were spun down at 1000g for 5 minutes to pellet the mucus in samples with a supernatant isolated for downstream analysis and kept frozen at -80°C until use. Saliva cytokine levels were quantified using Human Cytokine Array / Chemokine Array 71-plex (Eve Technologies, Calgary, Alberta, Canada).

### Bioinformatic Processing and Statistical Analysis

For microarray datasets, raw intensity values and annotation data were downloaded. Probe definition files for the Clariom D human microarray chip were retrieved from Bioconductor and probes were annotated with Ensembl IDs in R (version 4.1.0). All gene expression data from a given study were unified into a single data matrix that was then normalized by robust multiarray average (RMA) normalization. Normalized expression levels for *CXCL10, CXCL11,* and *TNFSF10* were extracted.

For RNA-seq datasets, raw count values and annotation data were downloaded directly from the corresponding author’s GitHub. Counts were normalized using the limma package in R (version 3.14) with the design matrix including virus, sex, and age annotation data. Normalized expression levels for *CXCL10, CXCL11,* and *TNFSF10* were extracted. Differential expression analysis was performed using the DESeq2 package in R (version 3.14).

Gene expression levels were tested for significant differences between viral infections or other phenotypic data of interest via pairwise Student’s t-tests with Benjamini–Hochberg multiple testing correction or via ANOVA followed by Tukey Honest Significant Difference *post-hoc* tests using the stats R package (version 3.6.2). Gene expression box plots were generated with GraphPad Prism 9 (GraphPad Software Inc., USA) and heat maps were generated using pheatmap R package (version 1.0.12). For heat maps, normalized expression levels are scaled by gene. Gene expression box plots were expressed as mean and standard errors of the mean (SEM) with unpaired t-tests performed to compare the means of two groups. Where three groups were compared, a one-way ANOVA was performed with a Bonferroni correction for multiple comparisons. To determine variance in *CXCL10, CXCL11*, and *TNFSF10* gene expression over time with SARS-CoV-2 infection, the host gene of interest was expressed as percent change from first period of inpatient sampling (set as time = 0) for each individual patient. Line graphs and box plots were generated with GraphPad Prism 9 (GraphPad Software Inc., USA).

Differences were considered statistically significant when p < 0.05.

To assess protein levels in healthy human saliva, quantitative proteomic data corresponding to ProteomeXchange dataset PXD003028 [48] were downloaded from the MaxQB Database [49]. Reverse hits, common contaminants including keratins, and low confidence hits were removed, and data consistency verified using Pearson’s correlation analysis within the Perseus platform [50]. Label free quantification intensities were subsequently summed to give a robust read-out for average protein levels for the 5551 proteins quantified. More than 95% proteins were identified in at least 3 samples, and nearly 80% across all 8 samples.

### CXCL10 LFA Prototype Development

#### Antibody pair selection process

Commercially available antibodies and reagents were selected for assay development to ensure reproducibility and robustness between labs. Antibody pairs for detection of CXCL10 were identified from commercially available sources that were amenable to ELISA applications. A recombinant anti-human CXCL10 mouse IgG-monoclonal (MAB2661), anti-mouse IgG goat-polyclonal (AF-266-NA), and recombinant CXCL0 protein (Product 266-IP) were selected from R&D Systems (Toronto, Ontario, Canada).

#### Conjugation of monoclonal CXCL10 antibody to 40nm gold

Conjugation was performed by using an N-Hydroxysuccinimide (NHS) coupling kit from Cytodiagnostics following manufacturer directions (Product CGN5K-40-2, Burlington, Ontario, Canada). Briefly, the monoclonal CXCL10 antibody was rehydrated in the supplied Protein Resuspension Buffer to a final concentration of 0.5 mg/mL. A single conjugation reaction was performed by addition of the supplied Reaction Buffer and CXCL10 solution to the lyophilized NHS-activated gold nanoparticle pellet. The reaction was gently mixed for 1 hr before adding 10 μL of the supplied Quencher Solution followed by 10 μL of 10% bovine serum albumin dissolved in distilled water. The 40 nm gold conjugate was pelleted by centrifugation (1300x g); the supernatant was removed, and 1 mL of Conjugate Buffer was added to resuspend and wash the conjugate before re-centrifugation to pellet the product. After removing the supernatant, the product was resuspended, and the OD adjusted to 10 using the provided Conjugate Buffer.

#### Preparation of lateral flow dipsticks

A control line solution (150uL) was prepared using 0.2 mg/mL of the anti-mouse IgG antibody reconstituted in a capture buffer (10 mM HEPES, pH 7.4, 0.1% BSA, 0.5% Methanol). The test line solution (150uL) was prepared with a polyclonal CXCL10 antibody at 0.1 mg/mL in capture buffer. Hi-Flow^TM^ Plus 135 (Catalog #HF135MC100, 60mm x 301 mm) nitrocellulose membranes were purchased from EMD Millipore Corporation, Burlington Massachusetts. The test and control solutions were striped in parallel across the non-pre-treated nitrocellulose membranes using a syringe pump set to 2.2mL/min connected to Claremont Bio’s Automated Lateral Flow Reagent Dispenser set at 3V. The resulting membranes were allowed to fully dry at room temperature for 18 hours before assembly with the wicking pad (Cellulose fibre sample pad, product #CFSP223000, EMD Millipore; Billerica, Massachusetts, USA) and cut into 4 mm wide strips by a CM5000 Guillotine Cutter. Batches of approximately 150 completed LF strips were made and stored at room temperature for up to 6 months without loss of sensitivity (data not shown).

#### Lateral flow sensitivity testing

Solutions for LFA sensitivity testing were prepared with 90% (by volume) lateral flow buffer (10mM HEPES, 150 mM NaCl, 0.1% Tween-20, 1% BSA, and 0.5% PEG 8000) and 10% (1 OD final concentration) monoclonal anti-human CXCL10 antibody 40nm gold conjugate. Standards of known concentration were generated by adding recombinant CXCL10 to Reconstitution Buffer (1X PBS pH 7.4, 0.1% bovine serum albumin) to generate 50, 20, 10, 5, and 2 ng/mL final concentrations along with a 0 ng/mL negative control.

Solutions for LFA testing with artificial saliva (product #1700-0316, ASTM E2721-16 with Mucin, pH 7.0; Pickering Laboratories, Mountain View, California, USA) were prepared with 45% (by volume) lateral flow buffer and 45% artificial saliva. Monoclonal anti-human CXCL10 antibody 40nm gold conjugate was added to a final concentration of 1 OD (or 10% by volume). Standards of known concentration were generated by adding recombinant CXCL10 to Reconstitution Buffer (1X PBS pH 7.4, 0.1% BSA) to generate 400, 50, 20, 10 and 5 ng/mL final concentrations along with a 0 ng/mL negative control.

For sensitivity studies in ideal buffer or artificial saliva, five unique replicates of standards were generated with 50ul of the solutions added to a 96-well plate. The solution was allowed to flow completely through the dipstick for 15 min and imaged with band intensity quantification performed using an IUL IPeak lateral flow strip reader (Product # 100033000; Barcelona, Spain) programmed to identify test and control bands.

#### Testing of LFA with human saliva from healthy subjects

Saliva was collected from healthy subjects with no symptoms of upper or lower respiratory tract infections. Subjects were asked to wash their mouth with water and discard into a sink, followed by 5 minutes of collection of 5 ml of collection (whichever came first). Samples were analyzed as is without centrifugation to represent a real-world point of care setting. Aliquots from 7 individuals were generated to create neat samples and CXCL10 spiked samples (10ng/mL). LFAs were completed by adding 50ul of sample to a 96 well plate followed by insertion of the dipstick for 15 minutes with qualitative visualization and photo capture completed.

## Results

To first investigate the potential association between *CXCL10*, *CXCL11*, and *TNFSF10* with viral infection, pre-processed transcriptomic datasets were examined using the Harmonizome resource (https://maayanlab.cloud/Harmonizome/)[45]. 366 processed microarray datasets under the category “GEO Signatures of Differentially Expressed Genes for Viral Infections”[51,52] were curated and filtered to include only datasets from experiments that used respiratory viruses and *in-vitro* human samples (**Figure 1A**). This filtering reduced the total number of GSE studies to 17 and corresponding microarray datasets to 199. From the 17 different studies, two were from a single time-point and the remaining 15 of the studies were time-series data. A variety of respiratory tract viral pathogens were used in the datasets including influenza A, SARS-CoV variants, human metapneumovirus, measles, RSV and RV (**See Supplementary Table 1**). The Harmonizome database extracted 600 differentially expressed genes following viral infection for each of the 199 microarray datasets which included the top 300 genes with increased expression and bottom 300 genes with decreased expression. Of the 199 datasets, there were more datasets with upregulation of *CXCL10, CXCL11*, and *TNFSF10* gene transcript relative to datasets where these genes were downregulated. *CXCL10* was found to be upregulated (Harmonizome association value > 1) in 39 datasets and downregulated (Harmonizome association value < -1) in 1 (**Figure 1B**). *CXCL11* was upregulated in 33 datasets. *TNFSF10* was upregulated in 36 datasets and downregulated in 3. Among the three genes, *CXCL10* was upregulated in the greatest number of viral infection datasets. The common housekeeping genes *GAPDH, TUBB* and *ACTB* were analyzed in all 199 datasets to provide context for the observed upregulation of *CXCL10, CXCL11*, and *TNFSF10*. (**Figure 1C**). The three housekeeping genes showed altered expression in fewer datasets than for *CXCL10, CXCL11*, and *TNFSF10*. These results provide evidence that *CXCL10, CXCL11*, and *TNFSF10* are upregulated agnostic to human cell type and pathogen type when filtered for human respiratory tract viral infections and provided a rationale to explore the candidates further with samples of patients with clinically diagnosed respiratory viral infections.

**Figure 1:**
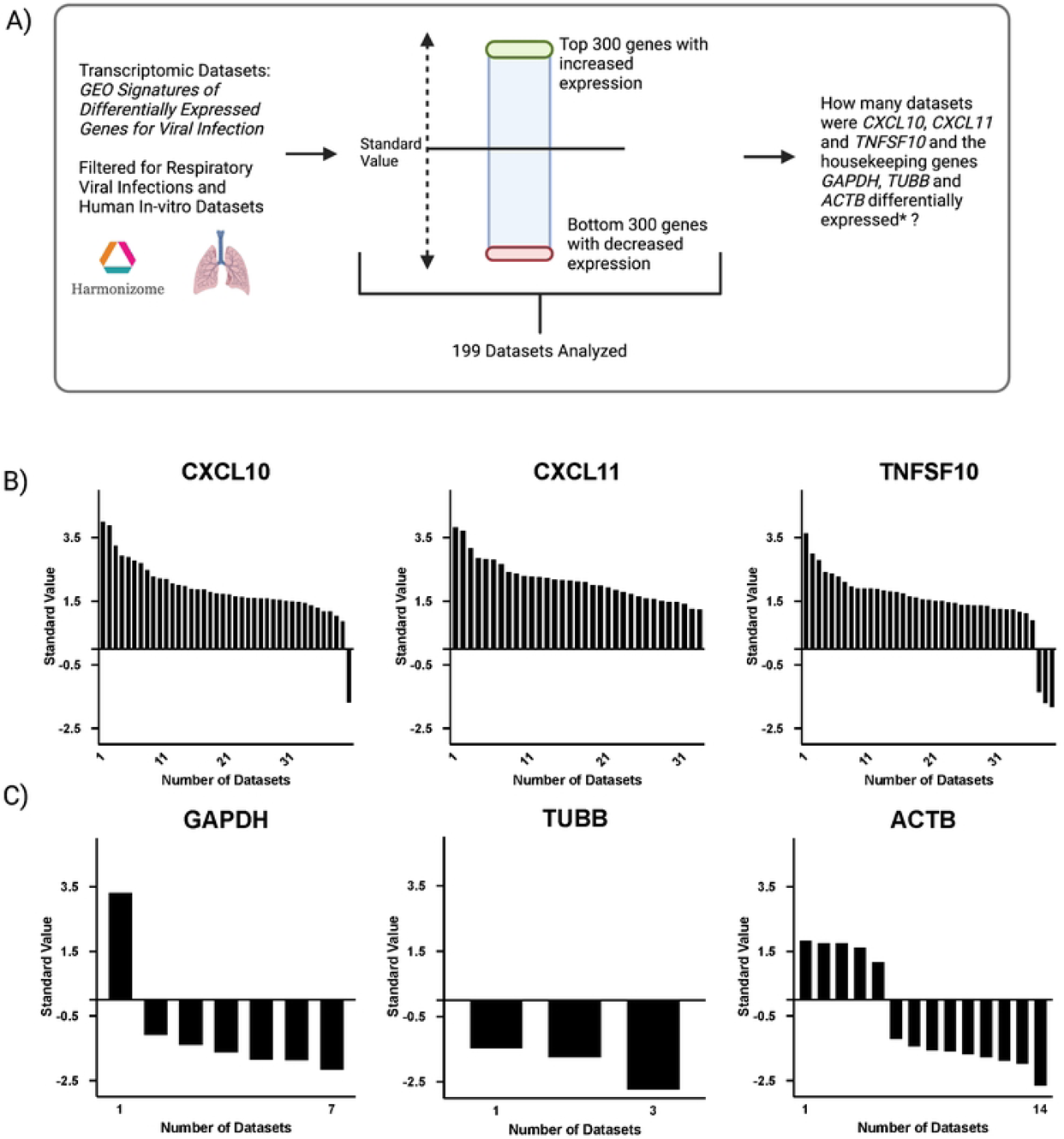
*CXCL10*, *CXCL11* and *TNFSF10* show increased expression *in-vitro* after viral infection. **A:** Schematic of workflow. Pre-processed transcriptomic datasets using the Harmonizome resource (Rouillard *et al*., 2006) database of processed microarray datasets under the category “GEO Signatures of Differentially Expressed Genes for Viral Infections” (Edgar *et al*., 2002; Barrett *et al*., 2013) was accessed for hypothesis testing for elevations in *CXCL10*, *CXCL11*, and *TNFSF10* following viral exposure *in vitro* from a variety of experimental conditions. This dataset contained 366 individual datasets of mRNA expression profiles using microarray technology. All non-human experiments and non-respiratory viruses were excluded which filtered down to 199 microarray datasets. The studies in which *CXCL10, CXCL11* and *TNFSF10* appeared to be differentially expressed were counted (*Found in the top 300 or bottom 300 differentially expressed genes with a Harmonizome standard value greater than 1 (up-regulated) or below -1 (down-regulated).) Datasets included but not limited to Calu-3 cell lines, HAE cultures infected with respiratory viruses including but not limited to FluA, SARS-CoV and Human metapneumovirus. **B:** *CXCL10* was found to be upregulated in 39 independent viral infection datasets, and downregulated in one. *CXCL11* was upregulated in 33 viral infection datasets. *TNFSF10* was upregulated in 36 and downregulated in 3. **C:** The housekeeping genes *GAPDH*, *TUBB* and *ACTB* were analyzed in all 199 and showed different expression levels in fewer datasets than for *CXCL10*, *CXCL11*, and *TNFSF10*. *GADPH* was found to be upregulated in 1 independent viral infection datasets, and downregulated in 6. *TUBB* was down in 3 viral infection datasets. *ACTB* was upregulated in 5 and downregulated in 9.

To interrogate the expression of *CXCL10*, *CXCL11*, and *TNFSF10* in clinical samples, we analyzed a combination of publicly available and in-house datasets from upper airway samples taken during studies of respiratory tract infections that included RSV, RV, influenza A and SARS-CoV-2 with corresponding non-infected negative controls (**Figure 2A**). *CXCL10* and *CXCL11* were upregulated with RSV infection relative to non-infected controls (**Figure 2B** – orange bars, p < 0.05) and to samples from RV infected subjects (p < 0.05). *CXCL10*, *CXCL11* and *TNFSF10* were not differentially expression between NPS samples collected from RV infected subjects relative to non-infected controls (**Figure 2B** – blue bars, p > 0.05). No significant difference was found between NPS samples from influenza A infected individuals relative to non-infected controls for *CXCL10*, *CXCL11* and *TNFSF10* (**Figure 2C**, p > 0.05).

**Figure 2.**
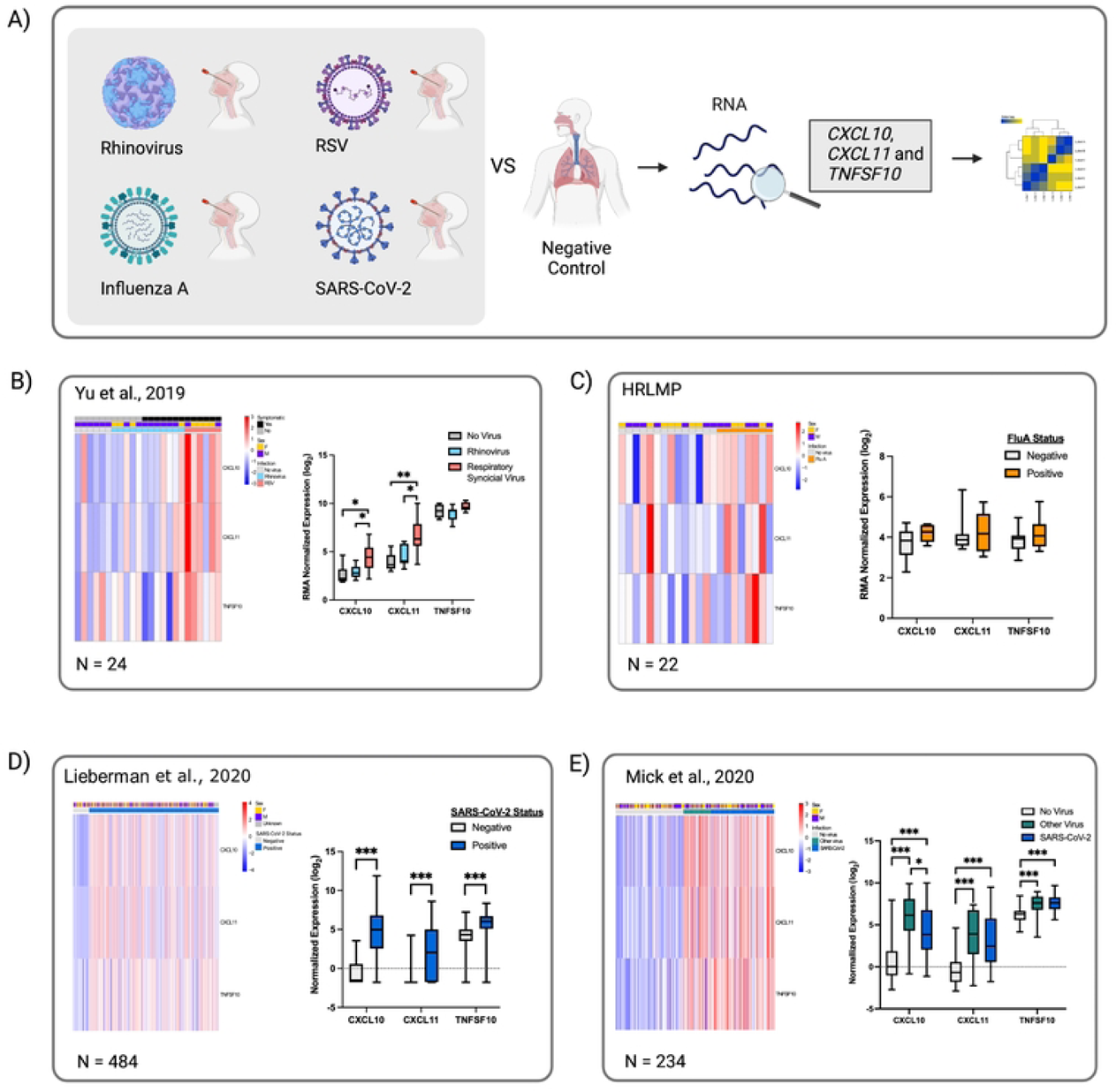
*CXCL10*, *CXCL11* and *TNFSF10* expression in nasopharyngeal swab and nasal mid-turbinate samples from subjects with different viral respiratory tract infections including SARS-Cov-2, FluA, Respiratory Syncytial Virus, Rhinovirus. **A:** Schematic of workflow. **B:** From Yu *et al*., 2019 - GSE117827: *CXCL10*, *CXCL11* and *TNFSF10 gene expression* was compared from the mid-turbinate nasal swab of pediatric subjects with RSV, RV and negative controls. Clustered heatmap of log_2_ expression levels annotated by symptomatic, sex and infection with blue representing decreased expression and red increased expression. On the right, boxplot of RMA normalized expression (log_2_) (n = 24). There was no significant difference when comparing rhinovirus infected NPS to healthy control gene expression for *CXCL10*, *CXCL11* and *TNFSF10* (p > 0.05). *CXCL10* and *CXCL11* up-regulation was positively correlated with Respiratory Syncytial Virus (RSV) when compared to healthy control (p = 0.016, p = 0.006). **C:** Hamilton Regional Laboratory Medicine Program (HRLMP) microarray data: *CXCL10*, *CXCL11* and *TNFSF10 gene expression* was compared from the NPS of subjects with influenza A (FluA) and negative controls. Clustered heatmap of log_2_ expression levels annotated by sex and infection status with blue representing decreased expression and red increased expression. On the right, boxplot of RMA normalized expression (log_2_) (n = 22). No significant difference between *CXCL10*, *CXCL11* and *TNFSF10* expression in FluA infection compared to negative control (p > 0.05). **D:** From Lieberman *et al*., 2020 - GSE152075: *CXCL10*, *CXCL11* and *TNFSF10 gene expression* was compared from the NPS of individuals with suspected SARS-CoV-2 infection. Clustered heatmap of log_2_ expression levels annotated by sex and infection status with blue representing decreased expression and red increased expression. On the right, boxplot of RMA normalized expression (log_2_) (n = 484). *CXCL10*, *CXCL11* and *TNFSF10* expression was significantly upregulated SARS-Cov-2 infection compared to those who tested negative (p < 0.001). **E:** From Mick *et al*., 2020 - GSE156063: *CXCL10*, *CXCL11* and *TNFSF10 gene expression* was compared from the NPS of subjects SARS-CoV-2 positive, SARS-CoV-2 negative but positive for another respiratory virus and no respiratory virus detected by metagenomic next generation sequencing (i.e., non-viral ARI such as bacterial infection). Clustered heatmap of log_2_ expression levels annotated by sex and infection status with blue representing decreased expression and red increased expression. On the right, boxplot of RMA normalized expression (log_2_) (n = 234). *CXCL10*, *CXCL11* and *TNFSF10* expression was significantly upregulated SARS-Cov-2 infection compared to healthy control (p < 0.001). * = p < 0.05, ** p < 0.01 and *** = p < 0.001.

With the COVID-19 pandemic, a variety of studies from SARS-CoV-2 infected subjects and non-infected controls have become publicly available. Probing a dataset by Lieberman *et al*., 2020, we observed increases in *CXCL10*, *CXCL11* and *TNFSF10* in upper airway samples from SARS-CoV-2 infected subjects compared to non-infected negative controls (**Figure 2D**, p < 0.001). Similarly, analyzing data from Mick *et al*., 2020, we observed increases in *CXCL10*, *CXCL11* and *TNFSF10* in samples from SARS-CoV-2 infected subjects compared to non-infected negative controls (**Figure 2E**, p < 0.001). Collectively, these data from multiple respiratory tract viral infections suggest that *CXCL10*, *CXCL11* and *TNFSF10* positively correlate with infection status, with a robust and validated increase observed during SARS-CoV-2 infection.

The observation that elevations in *CXCL10*, *CXCL11* and *TNFSF10* transcripts from upper airway swab samples strongly associated with COVID-19 infection, prompted further exploration with additional SARS-CoV-2 study samples with data features of viral load and mortality (**Figure 3A**). Using a dataset from 735 subjects (519 PCR confirmed SARS-CoV-2 negative and 216 PCR confirmed SARS-CoV-2 positive) that included metadata on Ct cycle for indication of viral load, we observed that *CXCL10*, *CXCL11* and *TNFSF10* gene expression showed trends for increasing with greater SARS-CoV-2 viral load quantified by PCR Ct value (**Figure 3B**). Pooling all SARS-CoV-2 positive samples together and comparing to SARS-CoV-2 negative samples, *CXCL10* showed an upregulation of log_2_ fold change of 3.4 (adjusted p value (q value) = 7.38E-31), *CXCL11* showed an upregulation of 2.9 (q value = 4.43E-22) and *TNFSF10* showed an upregulation of 1.0 (q value = 9.39E-23). Conversion of SARS-CoV-2 RT-PCR cycle threshold (Ct) values into a continuous variable showed highly significant correlations between this measure of viral copy number and the three candidate biomarkers (*CXCL10* - q value = 1.23E-54, *CXCL11* - q value = 5.17 E-47, and *TNFSF10* - q value = 4.26E-38). Individuals who tested positive for other respiratory viral infections, under subclass “Other viral infection” appeared to also correlate with higher expression of *CXCL10*, *CXCL11* and *TNFSF10* (**Figure 3B –** pink bar for metadata on virus subclass), although no metadata on the other infection status was available. As viral load may not be an accurate indicator of infection severity in COVID-19 [53,54], we quantified *CXCL10*, *CXCL11* and *TNFSF10* transcript levels in fatal COVID-19 inpatients via microarray gene chip. Using a dataset of 25 subjects (19 PCR confirmed SARS-CoV-2 positive survivors and 6 PCR confirmed SARS-CoV-2 positive deaths) the mean RMA gene expression value for fatal vs non-fatal COVID cases was 4.3 (SD 1.8) vs 3.5 (SD 1.11) for *CXCL10*, 3.7 (SD 0.4) vs 3.3 (SD 0.4) for *CXCL11* and 6.0 (SD 1.5) vs 6.3 (SD 0.8) for *TNFSF10*. In this limited sample size, mean values were higher in COVID-19 fatal cases for *CXCL10* and *CXCL11* but were not significantly different (**Figure 3C**, p > 0.05). No trends were observed for *TNFSF10*.

**Figure 3:**
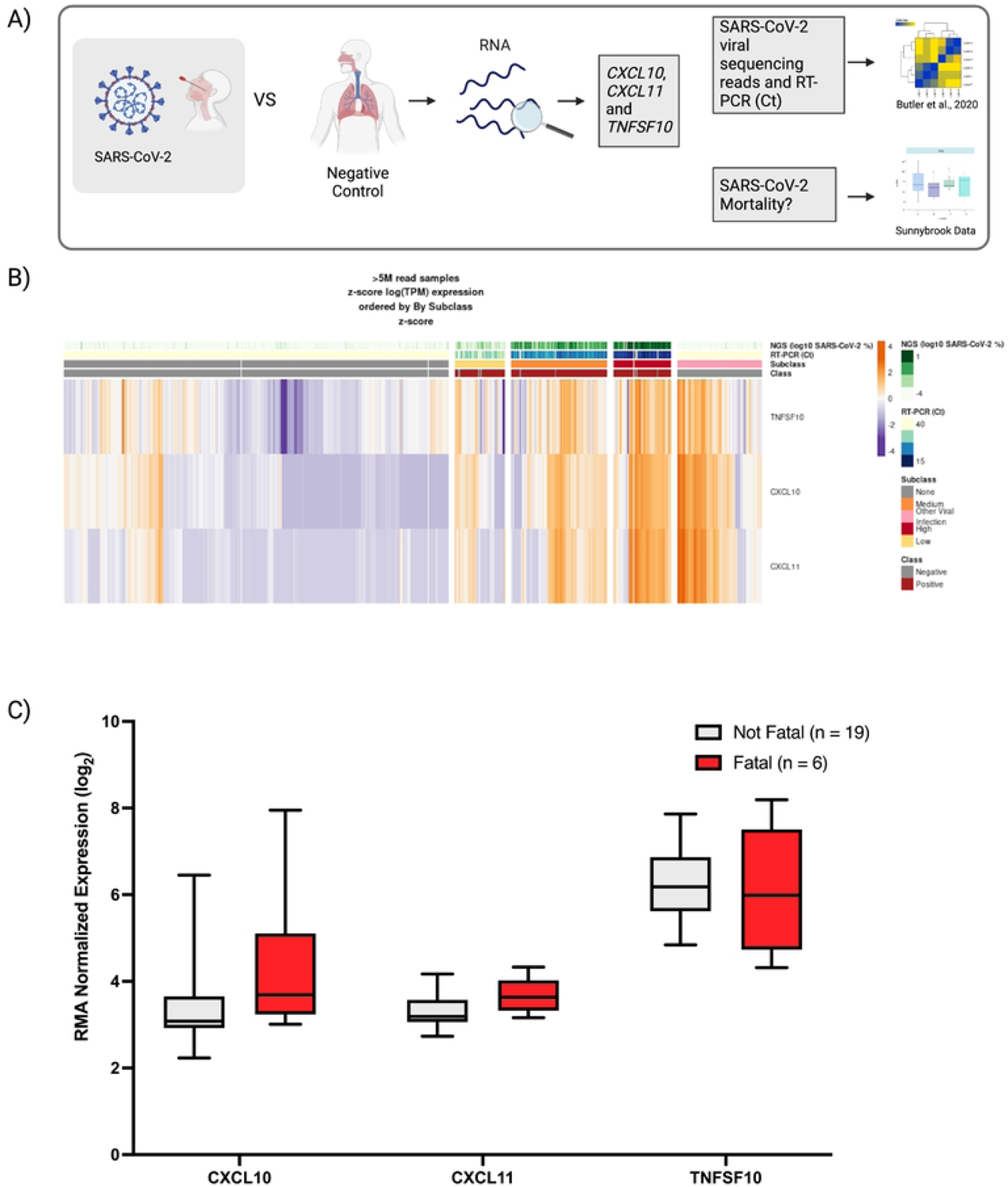
The *CXCL10/CXCL11/TNFSF10* gene signature in nasopharyngeal swab samples is positively correlated with SARS-CoV-2 viral load. **A:** Schematic of workflow. **B**: SARS-CoV-2 viral sequencing reads and qPCR cycle thresholds correlate with the *CXCL10/CXCL11/TNFSF10* gene signature. (N = 735). The “Viral Level Continuous” comparison group converted qRT-PCR cycle threshold (Ct) values into a continuous variable by inverting CT values where Ct = 15 is equal to 1.0 and a Ct > 40 is 0, *CXCL10* showed an upregulation of log_2_ fold change of 6.2 (q value = 1.23E-54), *CXCL11* showed an upregulation of log_2_ fold change of 6.0 (q value = 5.17 E-47) and *TNFSF10* showed an upregulation of log_2_ fold change of 1.9 (q value = 4.26E-38). Data and Figure from Butler *et al*. 2021 - For research purposes only. All rights reserved. © Mason Lab and Weill Cornell Medicine, 2020). **C:** Mortality of COVID-19 patients is associated with only modest changes in the *CXCL10/CXCL11/TNFSF10* gene signature at the time of original patient sampling. No significant correlation was found (p > 0.05).

A limitation of leveraging swab samples at the time of diagnosis and examining host responses is that all samples are not collected at the same time during the course of infection. As host responses and antiviral responses will vary throughout an infection, we set out to quantify *CXCL10, CXCL11,* and *TNFSF10* gene expression and variance over the course of SARS-CoV-2 infection by serial NPS sampling over the period of hospitalization. Using an inpatient cohort of 6 subjects (5 females, 1 male – 72.8 years mean age +/-13.3 years), serial samples were collected when study subjects felt well enough to provide a sample (**Figure 4A**). *CXCL10*, *CXCL11* and *TNFSF10* transcripts were quantified in all samples provided until the patient was discharged from the inpatient unit (range from 12 to 26 days). Variance for *CXCL10* was 0.213, *CXCL11* was 0.172, and *TNFSF10* was 1.679 (**Figure 4B-C**). The variance for each gene was calculated per patient and averaged for each gene (grey bars), with statistically lower variance observed for *CXCL10* and *CXCL11*, with both genes having lower variance than *TNFSF10* (p<0.05). The data from the studies analyzed thus far suggest that *CXCL10* and *CXCL11* are elevated with SARS-CoV-2 infection, track with viral copy number, and are stable throughout the progression of COVID-19 infection. These data suggest that CXCL10 and CXCL11 may be relevant biomarkers that could be useful for tracking respiratory tract viral infections.

**Figure 4:**
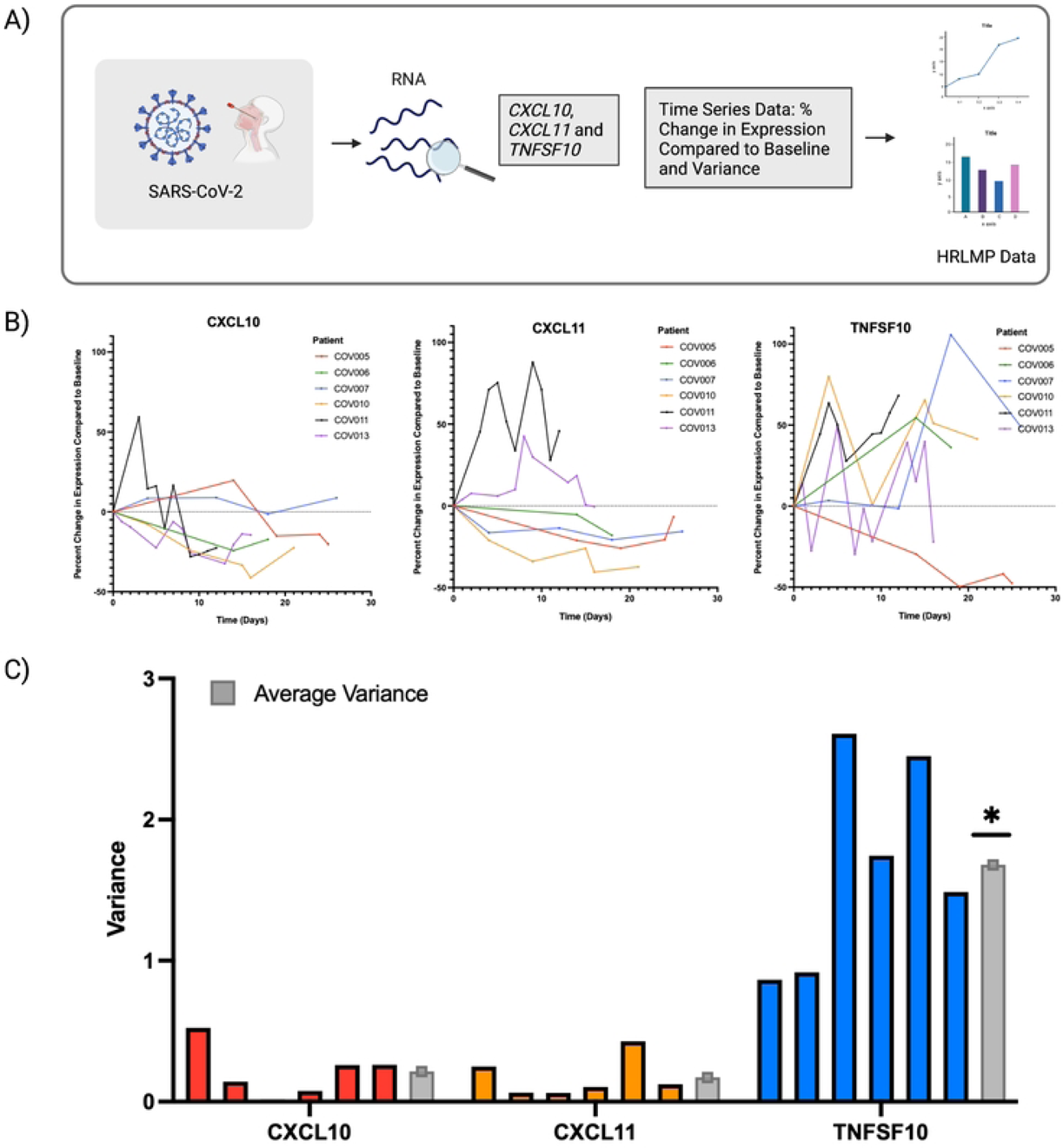
*CXCL10/CXCL11/TNFSF10* gene signature expression over the course of COVID-19 infection as measured by serial nasopharyngeal swab sampling over period of hospitalization. **A:** Schematic of workflow. **B:** *CXCL10*, *CXCL11*, and *TNFSF10* expression over time in hospitalized COVID-19 positive patients expressed as % change from first sampling (set as time=0). The data was collected from 6 independent patients (COVXXX) who had distinct sampling counts dependent on clinical management of COVID-19 infection. COV005 = 5 measurements, COV006 = 3 measurements, COV007 = 5 measurements, COV010 = 6 measurements, COV011 = 10 measurements, COV013 = 11 measurements. **C:** To determine which gene fluctuated the least of the course sampling, the variance of RMA values for *CXCL10/CXCL11/TNFSF10* were calculated for each patient (colours) and averaged (grey). *CXCL11* variance = 0.17, *CXCL10* variance = 0.21 and *TNFSF10* variance = 1.68. * p < 0.05 relative to mean variance for *CXCL10* and *CXCL11*.

The data presented identify that *CXCL10* gene demonstrates the greatest magnitude of change during SARS-COV-2 infection in upper airway samples collected via NPS. NPS are not routinely self-administered and present a problematic sampling site for at-home or healthcare resource deficient settings, which has preference to shallow nasal swabs or oral swabs or combination[55]. NPS collected during viral screening are frequently stored in transport medias that are optimized for nucleic acid stabilization and isolation relative to protein stability[56]. Oral sampling has emerged during the COVID-19 pandemic as a useful surrogate for the upper respiratory tract that can be performed by non-healthcare professionals at a point of care/need that is amenable to both nucleic acid or protein detection strategies [57]. We therefore explored the utility of detecting CXCL10 protein in oral sampling using saliva collections as a sample format. To be a useful biomarker for monitoring respiratory tract infections, CXCL10 protein should not be detectable in healthy subject samples and only appear once a clinically meaningful infection occurs. Using a mass spectrometry-based proteomics dataset from 8 subjects (4 males and 4 females) that were sampled immediately after waking and again after first meal, we probed the healthy saliva proteome for CXCL10 (**Figure 5A**). In a list of 5551 proteins identified by label free quantification (**Figure 5B** and **Supplement Table 2**) CXCL10 protein was not found at any ranking. A table of the 15 most abundant identified proteins lists amylase as the top hit, confirming our analysis pipeline (**Figure 5C**). We next quantified the expression of CXCL10 protein in the saliva from a cohort of PCR-confirmed COVID-19 inpatients and healthy uninfected controls using a human cytokine/chemokine biomarker assay (Eve Technologies). The mean concentration of CXCL10 was 86.4 pg/mL (SD = 109.6) in healthy saliva and 1186.6 pg/mL (SD = 1252.3) in COVID-19 inpatients (**Figure 5D**, p < 0.05).

**Figure 5:**
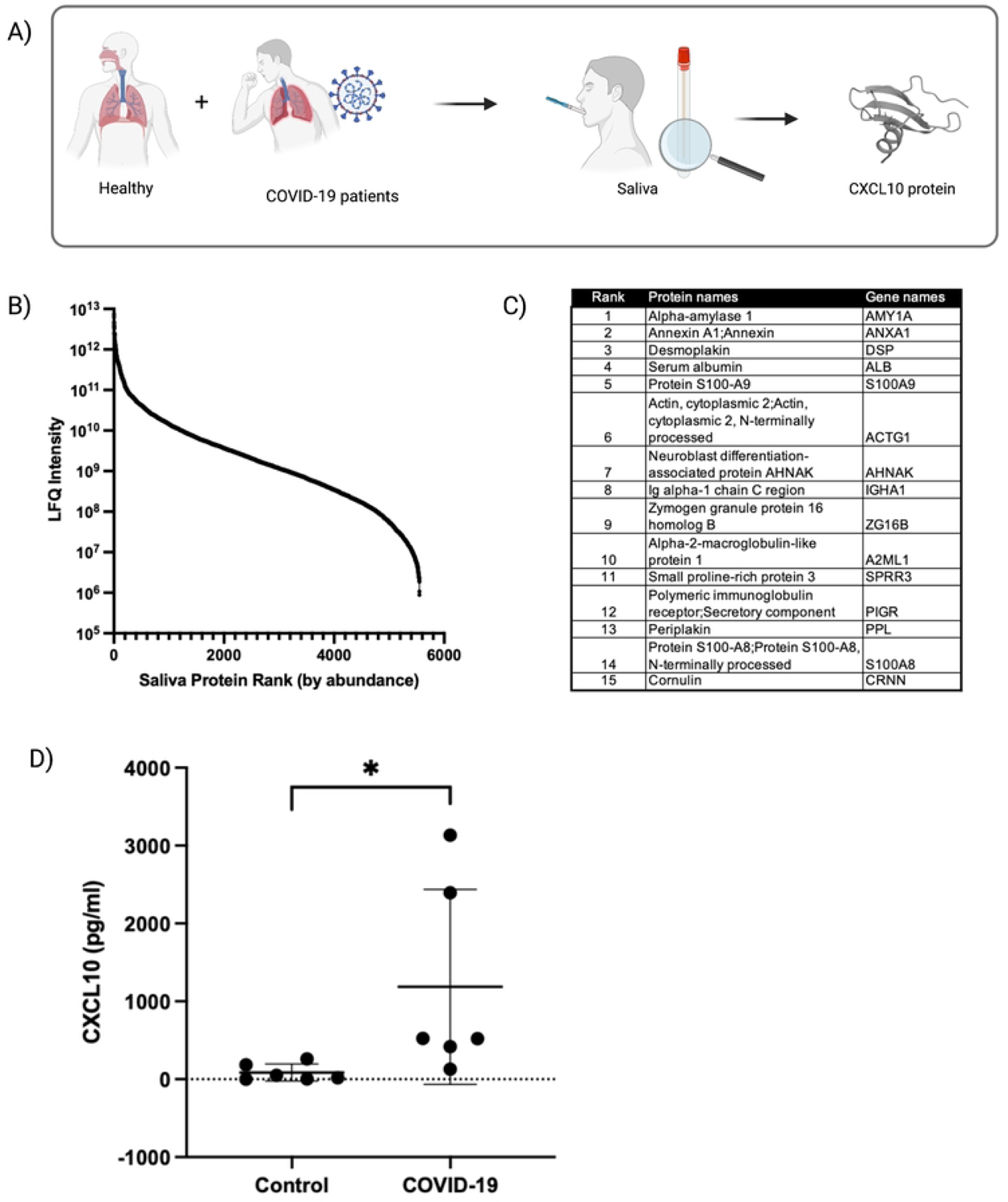
CXCL10 protein is not present in healthy saliva samples and elevated during infection in saliva from COVID-19 patients. **A:** Schematic of workflow. **B-C:** From Grassl *et al*., 2016 ultra deep analysis of the healthy saliva proteome, the CXCL10 protein was not found amongst the list of 5562 identified proteins (Supplementary Table 2). **D:** CXCL10 Levels in saliva of SARS-CoV-2 in hospitalized COVID-19 patients quantified using the Human Cytokine Array / Chemokine Array 71-plex (Eve Technologies, Calgary, Alberta, Canada). Healthy volunteers without symptoms of a respiratory infection were used as the control group. Mean concentration of CXCL10 in saliva was 86.4 pg/mL (SD = 109.6, n = 6) in healthy subjects, while a mean of 1186.6 pg/mL (SD = 1252.3) was observed in COVID-19 patients. The COVID-19 group showed a significantly greater CXCL10 concentration (* = p < 0.05).

The low levels of CXCL10 protein in saliva of healthy subjects but elevated levels in COVID-19 subjects is amenable to conventional LFA development that detects the upregulation of a mediator of interest. LFAs have become widely accepted tools for self-testing during the COVID-19 pandemic and represent a technology platform useful for at-home and healthcare deficient settings. Therefore, we next pursued the development of an open-source CXCL10 LFA using commercially available reagents (**Figure 6A**). A recombinant anti-human CXCL10 mouse IgG-monoclonal and anti-mouse IgG goat-polyclonal were selected as an antibody pair for CXCL10 detection. Assay development was validated in ideal buffer demonstrating a sensitivity of 2ng/mL (**Figure 6B-C**). Assay testing in commercially available artificial saliva reproduced the sensitivity of 2ng/mL (**Figure 6D-E**). Lastly, the assay detected 10ng/mL CXCL10 protein spiked into real human saliva from healthy controls that was not detected in control /non-spiked samples from the same donors (**Figure 6F**). These results demonstrate the sensitivity of an open-source CXCL10 LFA prototype for self-administered point of care saliva testing with potential applications in respiratory tract viral infection monitoring.

**Figure 6:**
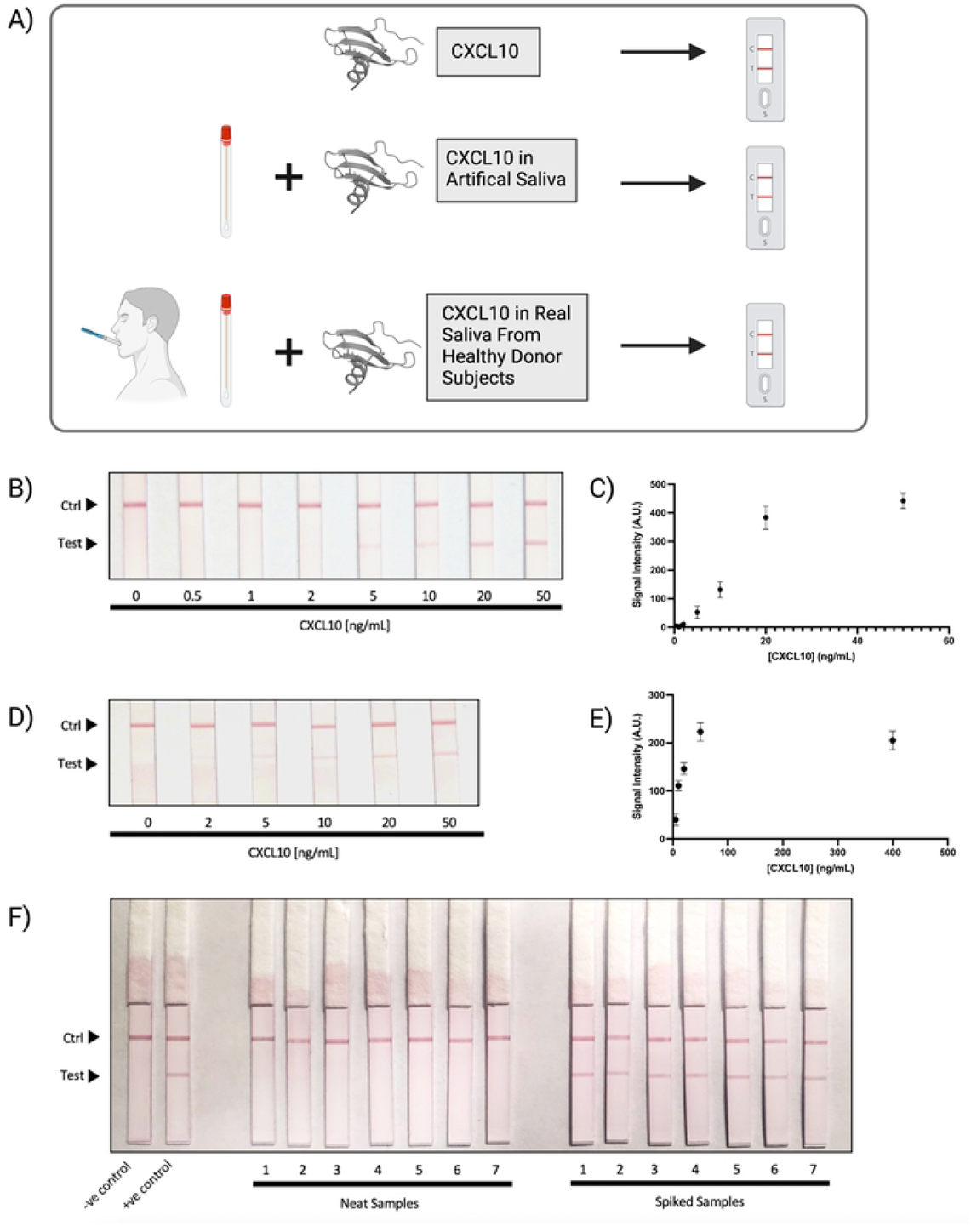
CXCL10 protein detection is feasible in fresh human saliva. **A:** Schematic of workflow. **B-C:** Sensitivity development and validation in ideal buffer (10mM HEPES, 150 mM NaCl, 0.1% Tween-20, 1% BSA, and 0.5% PEG 8000 – see methods for more details). Limit of detection of CXCL10 using a hand-held reader. A positive signal is generated at 2ng/mL with n = 5. **D-E:** Sensitivity test in artificial saliva (product #1700-0316, ASTM E2721-16 with Mucin, pH 7.0; Pickering Laboratories, Mountain View, California, USA) prepared with equal parts lateral flow buffer and artificial saliva - see methods for more details. **F:** Real world testing in human saliva from healthy control without (neat) and with (spiked) CXCL10 (10ng/mL) addition.

## Discussion

ISGs play a key role in defence against viral infections and measuring their expression levels during respiratory infections may provide diagnostic and prognostic information beyond measurement of a pathogen alone. In the present paper, we characterize three ISGs of interest that result in secreted protein products amenable to extracellular detection strategies: *CXCL10*, *CXCL11* and *TNFSF10.* We quantify *CXCL10*, *CXCL11* and *TNFSF10* expression levels in the context of multiple respiratory tract viral pathogens including RSV, RV, influenza A and SARS-CoV-2. Gene expression levels from upper airway samples suggested that *CXCL10* elevations were consistent across multiple viruses, had qualitative positive trends with measures of severity of infection (e.g., viral load), and had the lowest variance over course of COVID-19 infection of the three candidates examined. To transition from upper airway samples collected by a healthcare professional to a self-collected sample format, we next explored CXCL10 levels in oral samples. A published deep proteomic profiling of healthy human saliva suggested low to absent levels of CXCL10 protein in healthy subjects. In contrast, saliva from SARS-CoV-2 infected individuals resulted in elevations in CXCL10. Leveraging this relative binary behaviour of CXCL10 in healthy vs SARS-CoV-2 infected sample groups, we developed a prototype LFA for CXCL10 protein with a sensitivity of 2ng/mL in human saliva. Collectively, our work adds to the growing importance of examining host biomarkers during respiratory tract infections for diagnostic and prognostic value and demonstrates the feasibility of using self-administered qualitative LFA detection methods.

Our study used a combination of publicly available datasets and *de novo* generated datasets. Consequently, the results are heavily dependent on the annotation of the data provided for each study. Differences in study design and the definition of control groups may contribute to potential biases and limitations in the data. Commercially available or bespoke PCR-based respiratory panels were used in several of the studies to annotate positive and negative subjects in individuals suspected of respiratory tract viral infection. PCR-based detection strategies can diagnose several viruses simultaneously while having greater challenges for diagnosing bacterial respiratory infections [58]. The challenges with bacterial diagnostic decisions in the respiratory tract are rooted into the commensal nature of bacteria in the nasopharynx such as *Streptococcus pneumoniae* and *Haemophilus influenzae* [59,60]. For example, up to 10% of adults are carriers of *Streptococcus pneumoniae*, creating a challenge to determine whether this organism is causing pathology or is simply part of the individual’s microbiome [59]. The potential for viral and bacterial co-infections as well as the presence of complex commensal communities can create limitations in analyzing data when only the viral infection status is provided. Any underlying virus or bacterial respiratory tract infection would impact host gene expression patterns that may include *CXCL10, CXCL11*, and *TNFSF10*. Furthermore, studies focused only on a single virus may not report the outcomes for other viruses detected in a multi-plex PCR assay. The publicly available COVID-19 negative labelled samples were generated during routine public SARS-CoV-2 testing campaigns. As a result, although an individual may be labelled as SARS-CoV-2 negative, they may have had an underlying viral or bacterial infection not quantified and recorded. Although the incidence of influenza A was low during COVID-19 [61,62], there remained the possibility that other respiratory tract viral or bacterial infections were present in individuals that tested negative for SARS-CoV-2 [40]. In Lieberman *et al*., 2020, patients suspected of SARS-CoV-2 infection were confirmed by RT-PCR result while those testing negative became the controls although no confirmations for other viral or bacterial respiratory tract infections were performed. Mick *et al*., 2020 took a distinct approach by using metagenomic RNA-sequencing and separating their cohort into three groups – 1) *SARS-CoV-2 positive* - no other viral RTI 2) *SARS-CoV-2 negative* - positive for another virus and 3) *SARS-CoV-2 negative* - no other RTI viruses detected. Metagenomic RNA-sequencing can simultaneously identify host genes and microbial RNA and can allow for an unbiased approach to labelling control groups and create further transparency when sharing data publicly [63]. Metagenomic molecular diagnostic strategies have been optimized for upper airway samples and are likely to be useful strategies moving forward with host-pathogen diagnostic approaches and be informative for future pandemics [64].

A variety of publicly available datasets and databases exist and are increasing in accessibility. Notable examples of databases include the NCBI resources Gene Expression Omnibus, the Broad Institute’s Connectivity Map, and the Cancer Genome Atlas. The diversity of these databases has been consolidated in meta-databases to attempt to harmonize publicly available resources. Harmonizome is a publicly accessible meta-database of 112 datasets from 65 resources (https://maayanlab.cloud/Harmonizome/about). We leveraged the consolidation performed by Harmonizome to determine the expression of *CXCL10*, *CXCL11*, and *TNFSF10* across transcriptomic datasets defined by “GEO Signatures of Differentially Expressed Genes for Viral Infections”. Annotation of the datasets under this search term revealed some non-human datasets and datasets derived from non-respiratory tract viral infections, which were subsequently removed to maintain focus on host-biomarkers to respiratory tract infections. A limitation of the datasets curated by Harmonizome is that they are pre-clinical studies that include multiple different viruses from multiple different cell-lines in addition to different time-points post-infection. Despite the potential for extensive variation in experimental conditions between datasets, a significant signal for an increase in *CXCL10*, *CXCL11*, and *TNFSF10* was observed across the meta-database, without observing the same for three common housekeeping genes, *ACTB*, *GADPH* and *TUBB*. Although Harmonizome contains multiple datasets and databases, an absence of well-annotated samples from respiratory tract viral infections in humans required additional dataset generation and curation to extend and validate observations of *CXCL10*, *CXCL11*, and *TNFSF10* elevations.

NPS and mid-turbinate swabs are standard clinical sampling strategies for analysis of respiratory tract viral infections and are amenable to PCR and transcriptomic based readouts [55,64,65]. Historically, the small sample amounts collected from upper airway swabs have limited the ability to perform transcriptomic analysis which resulted in a focus on multi-plex PCR technologies [66]. The development of microarray technologies for small amounts of input material and the reduction in sequencing costs that affords greater sequencing depth have increased accessibility to transcriptomics. Prior to COVID-19 pandemic, relatively few studies of respiratory tract viral infections capitalized on technological advances for transcriptomics on upper airway samples. A seminal study demonstrating the ability to perform unbiased transcriptomics on upper airway samples was performed on a pediatric cohort of individuals with suspected and subsequent confirmation of respiratory tract infection via PCR [46]. We explored this publicly available host transcriptomic dataset from a cohort of 24 subjects. Independent of data that defines the time of initial infection, *CXCL10* and *CXCL11* up-regulation was positively correlated with RSV infection when compared to control. In contrast to RSV, no significant differences were observed for *CXCL10*, *CXCL11* and *TNFSF10* when comparing upper airway swabs from RV infected individuals to controls. The small sample size of this study precludes drawing conclusions about the absence of signal for our candidate host biomarkers with RV infection. Related to sample sizes of publicly available host-transcriptomic datasets from upper airway samples, the COVID-19 pandemic stimulated the mass adoption of molecular sequencing technologies and has since demonstrated the feasibility and utility of pursuing this approach, laying the foundation for a future state of host-pathogen diagnostics [35,40,47,64]. A limitation of leveraging clinical samples taken during diagnostic processes for research purposes is that there is lack of control over the time of initial infection, which could impact host gene expression signatures, particularly of ISGs which are time-sensitive [23]. This limitation may be overcome in the future with specific research study designs that could include prospectively following cohorts in a surveillance design to capture natural infections [66] or controlled human studies with viral challenges [67] with informed sample size calculations made based on the emerging publicly available datasets.

The presence of an elevated host biomarker during a confirmed respiratory tract viral infection may provide utility in determining stratifying those individuals at risk of morbidity and mortality, which could help inform treatment options at the time of first diagnosis [25,26]. To explore the possibility that either *CXCL10, CXCL11* and *TNFSF10* could function as a prognostic biomarker for disease severity, we explored expression levels in the context of SARS-CoV-2 viral copy number and COVID-19 mortality as datasets from other respiratory tract viral infections and viral copy are not publicly available. *CXCL10* and *CXCL11* have been shown to significantly increase in expression in NPS samples from SARS-CoV-2 patients compared with control[68]. Using a large publicly available host transcriptomic dataset with PCR confirmed SARS-CoV-2 positive cases and reported Ct values, thus *CXCL10, CXCL11* and *TNFSF10* are quantitatively observed to correlate with viral copy number (https://covidgenes.weill.cornell.edu/). The correlation between SARS-CoV-2 viral load in nasal samples tracking with severe disease has been supported in most but not all cases. In a prospective study, lower Ct values (i.e. higher viral load) correlated with severe disease in hospitalized patients. Although, there was no significant correlation between lower Ct values (i.e. higher viral load) for the risk of being hospitalized for SARS-COV-2 when adjusted for the time of symptoms onset [53]. A longitudinal study found increased SARS-CoV-2 viral load correlated with a higher risk of death, and increased inflammatory markers CRP and IL-6 [54]. Conversion of viral copy Ct value to a continuous variable (> 40 = 0, < 15 = 1) resulted in a statistically significant correlation between *CXCL10, CXCL11* and *TNFSF10* and amount of virus present. Despite the reports that SARS-CoV-2 viral copy number may relate to severity of COVID-19 disease, differences in the measure of severity and the populations chosen in independent studies have contributed to ambiguous conclusions. In addition to viral copy number, host responses have been measured throughout the COVID-19 pandemic and have been demonstrated to have predictive value for determining severity. Indeed, host responses to SARS-CoV-2 infection including serum IL-6 and C-Reactive Protein have been evaluated and demonstrated prognostic value in triaging COVID-19 patients [69,70]. In parallel to analyses relating viral copy number to host responses, we investigated death as an unambiguous measure of severity. In a cohort of 25 intensive care unit admitted subjects, each with PCR-confirmed SARS-CoV-2 infection, the upper airway swab sample taken at time of diagnosis showed no significant differences in magnitude of *CXCL10, CXCL11* and *TNFSF10* gene transcript between fatal and non-fatal COVID-19 cases. Previous research in blood samples of COVID-19 patients looked at 53 potential biomarkers and found CXCL10 as the best predictor of death [37]. They also observed that CXCL10 levels were increased in patients with ICU care compared to without ICU and CXCL10 levels decreased in COVID-19 survivors who were discharged from the hospital [37]. However, this study used a different sample type, blood, and they used a Bio-Plex Pro™ Human Cytokine 27-plex assay to determine CXCL10 protein concentrations. In addition to Lorè *et al*., 2021, several other studies using blood samples found CXCL10 was associated with COVID-19 disease severity [71,72]. These data suggest that focusing on blood as the sample type may provide a greater differentiating signature than upper airway gene transcripts. To date, point of care blood sampling for host biomarkers has received little traction in the infectious disease space. In contrast, self-administered LFAs for detection of protein in oral and upper airway samples provide a strategy for implementing point of care monitoring of host biomarkers relevant in respiratory tract infections.

The significant limitation of being blind to the time-point of initial infection is intrinsic to cross-sectional studies performed on individuals during initial diagnosis. To overcome this limitation, time-series studies may be performed to capture natural infections in the community, or the time course of infection can be monitored once an individual is admitted to the healthcare system, with admission becoming a baseline for the given study subject [66]. We took this secondary approach of defining baseline as the time of hospital admission and analyzed the variance in *CXCL10, CXCL11* and *TNFSF10* over the course of hospitalization. Analysis of variance for each biomarker in study subjects repeatedly sampled during their hospitalization was completed with each individual’s expression levels at admission functioning as their baseline. We quantified variance for each gene over time to identify which candidate may function as a more robust measure of host response to SARS-CoV-2 infection that is independent of the time of first sampling and diagnosis. The observation that *CXCL10* and *CXCL11* had lower variance over *TNFSF10* supported excluding the latter as a robust biomarker stable throughout the course of an infection. The stability of *CXCL10* and *CXCL11* that we observed in upper airway samples has been observed in serial blood and serum samples from COVID-19 patients [71,72]. Previous *in-vitro* studies found CXCL10 and CXCL11 expression in SARS-CoV-2 infected Calu-3 human lung cells peaked at 72 hours post infection [39]. CXCL10 showed greater log fold change and remained significantly elevated from 12-72 hours post-infection and CXCL11 elevated 18-72 hours post-infection [39]. Collectively, these data support the utility of CXCL10 protein as a prognostic biomarker for COVID-19 and provided a rationale for our focus on this mediator for downstream LFA development for self-administered testing.

Self-administered saliva testing for respiratory tract viral infections has become a reality through the COVID-19 pandemic [57]. The qualitative nature of LFA has been challenged in the popular and academic circles [73] with the net result being the widespread adoption of the technology on a global scale to provide a degree of diagnostic capacity previously not available to the general public. The possibility that other saliva-based tests relevant in infectious disease may emerge has therefore increased and may include host biomarkers of clinical importance [74]. The qualitative nature of LFAs requires a clear binary response of presence and absence for the test to be practical. Our *a priori* decision to characterize *CXCL10*, *CXCL11*, and *TNFSF10* were based on the assumption of their extracellular secretion with infection and low levels during times of no infection. In a deep proteomic profiling dataset of healthy human saliva from males and females with repeated measures, CXCL10 was not identified in a list of 5551 proteins detected, with 95% of the proteins identified in at least 3 samples and nearly 80% across all 8 samples. In contrast to deep proteomic profiling, we used a multiplex cytokine array from Eve Technologies and were able to detect CXCL10 with a mean concentration of 86.43 pg/mL (SD = 109.6, n = 6) in saliva from healthy subjects. Although these data may appear in conflict, an alternative explanation is that even deep proteomic profiling may not achieve the sensitivity of commercially available established multiplex cytokine arrays. Importantly, an elevation in CXCL10 protein was observed when analyzing saliva samples from COVID-19 infected subjects. The observed elevation of saliva CXCL10 protein provides a rationale to explore LFA technology for point of care detection and provides guidance on the sensitivity requirements for distinguishing samples from negative non-infected individuals and positive SARS-CoV-2 infected individuals. Leveraging commercially available reagents for LFA development including detection and capture antibodies, nitrocellulose membranes, and absorbent wicking material, we developed a prototype CXCL10 protein detection assay with a sensitivity of 2 ng/mL in artificial and healthy saliva. To our knowledge, this was the first CXCL10 LFA designed for human saliva samples. A previous study developed a CXCL10 LFA for amniotic fluid as a marker of intra-amniotic inflammation with sensitivity of 100 pg/mL (0.1 ng/mL)[75]. This study was able to achieve greater sensitivity, suggesting that the prototype presented could be optimized for detection of CXCL10 levels relevant during respiratory tract infections but not during periods of health. Importantly, if the sensitivity of a CXCL10 LFA were to be reduced to a level that can detect protein in healthy subjects, the utility of the test would be reduced as it would not help differentiate elevated levels of CXCL10 that may predict COVID-19 severity. Consistent with this requirement, CXCL10 protein levels peaked at around 60 ng/mL on day 2 of RV infection in adult nasal lavage samples which is within the range of our prototype CXCL10 LFA [33]. The concentration of CXCL10 measured with ELISA on the day of intubation from infants with RSV was observed to be around 33 ng/mL from bronchoalveolar lavage samples which is also within the range of our CXCL10 LFA [31].

Our characterization of CXCL10 gene expression in several datasets and development of a prototype CXCL10 responsive LFA amenable to saliva sampling represent first steps in a process towards biomarker validation and demonstration of clinical utility. Future utility could include examining the ability of CXCL10 protein levels in saliva to aid in discrimination of viral vs. bacterial respiratory tract infections, which would aid in rapid decision making for antibiotic administration as part of antibiotic stewardship practices. Future studies from clinically phenotyped subjects with respiratory tract bacterial infections in the absence of viral infections or bacterial/viral co-infections are required for this important research question to be answered. Perhaps closer to the clinic due could be the prognosis of COVID-19 during SARS-CoV-2 infection. Previous research has found CXCL10 to play a key role in predicting ICU admission using blood sample [37,71,72]. To validate CXCL10 in saliva as a biomarker of SARS-CoV-2 severity, further longitudinal research into time series data would be required. This would need to prove that CXCL10 levels are increased in a time frame where appropriate intervention is possible. It would not be useful to measure CXCL10 to predict severity if there are other more obvious clinical symptoms already present.

To conclude, we present data that characterizes *CXCL10*, *CXCL11* and *TNFSF10* gene expression in upper airway samples from individuals with respiratory tract viral infections. We provide a justification that CXCL10 protein product is amenable to LFA testing in a point of care setting. Further clinical validation of CXCL10 gene and protein as a biomarker of respiratory tract viral infections is required and should be complemented with study cohorts that include bacterial infections in the absence of viral infections. The utility of measuring CXCL10 at the point of care for diagnostic and prognostic purposes will grow as the appreciation that both host and pathogen molecular markers are important for optimal clinical care and healthcare system utilization.

## Data Availability

Table 1 provides details on the publicly available datasets that were used including URLs, gene expression method used, and notes on the samples from the datasets. Supplementary Table 1 provides information on the publicly available datasets that were retrieved from Harmonizome. In-house gene expression data analyzed with Clariom D microarray technology will be deposited to GEO upon acceptance with appropriate URL, accession numbers, and DOIs provided.

## Acknowledgements

The authors thank all of the authors of the publicly available datasets used who have facilitated analysis through email correspondence. We would also like to acknowledge Reina Ditta, Amanda Hodge, and Guillaume Pare from the Genomics and Molecular Epidemiology Lab at McMaster University for their core services for microarray gene expression assays. The authors also acknowledge Mark Loeb and Pardeep Singh for sample procurement and process optimization required for nasopharyngeal samples. BioRender was used for figure generation.

## Funding

This work was supported by the Roche Canada COVID Open Innovation Challenge, Ontario Government COVID-19 Rapid Research Fund, and the Thistledown Foundation/FastGrants.org. Hirota JA is supported by the Canada Research Chairs program and an Ontario Early Researcher Award. Doxey AC is supported by NSERC and an Ontario Early Researcher Award, and Eckhard U by research fellowships from the Beatriu de Pinós and Ramon y Cajal programs.

## Notes

### Competing Interest Statement

The authors have declared no competing interest.

### Funding Statement

Yes

### Author Declarations

For any human sample analysis the Hamilton Integrated Research Ethics Board (HiREB) provided review and approval of the study. Procurement of nasopharyngeal swab (NPS) and saliva samples from consented study subjects was approved under HiREB protocols 4914T, 5099T, and 10771.

